# The HUNT Study: a population-based cohort for genetic research

**DOI:** 10.1101/2021.12.23.21268305

**Authors:** Ben M. Brumpton, Sarah Graham, Ida Surakka, Anne Heidi Skogholt, Mari Løset, Lars G. Fritsche, Brooke Wolford, Wei Zhou, Jonas Bille Nielsen, Oddgeir L. Holmen, Maiken E. Gabrielsen, Laurent Thomas, Laxmi Bhatta, Humaira Rasheed, He Zhang, Hyun Min Kang, Whitney Hornsby, Marta R. Moksnes, Eivind Coward, Mads Melbye, Guro F. Giskeødegård, Jørn Fenstad, Steinar Krokstad, Marit Næss, Arnulf Langhammer, Michael Boehnke, Gonçalo R. Abecasis, Bjørn Olav Åsvold, Kristian Hveem, Cristen J. Willer

## Abstract

The Trøndelag Health Study (HUNT) is a population-based cohort of ∼229,000 individuals recruited in four waves beginning in 1984 in Trøndelag County, Norway. ∼88,000 of these individuals have available genetic data from array genotyping. HUNT participants were recruited during 4 community-based recruitment waves and provided information on health-related behaviors, self-reported diagnoses, family history of disease, and underwent physical examinations. Linkage via the Norwegian personal identification number integrates digitized health care information from doctor visits and national health registries including death, cancer and prescription registries. Genome-wide association studies of HUNT participants have provided insights into the mechanism of cardiovascular, metabolic, osteoporotic and liver-related diseases, among others. Unique features of this cohort that facilitate research include nearly 40 years of longitudinal follow-up in a motivated and well-educated population, family data, comprehensive phenotyping, and broad availability of DNA, RNA, urine, fecal, plasma, and serum samples.

## Background

Norway, like other Nordic countries, has characteristics that are uniquely favorable for recruitment to population studies, establishing biobanks, and identifying clinical outcomes and prospective disease trajectories. This includes a unique personal identification number applied throughout the life span, a universal and digitized public health care system, and accessible harmonized electronic health records. In addition, seventeen mandatory and validated national health registries are used for health analysis, administration, and emergency preparedness, and fifty-two national medical quality registries provide disease specific data on diagnosis and treatment parameters. Finally, Norwegians are an altruistic, highly motivated population for participating in biomedical research, as reflected in survey response rates of up to 89%. These factors have supported the establishment and maintenance of the Trøndelag Health Study (HUNT), a large population-based prospective Norwegian cohort, linked to registries and biobanks dating back more than 50 years (**Figure 1**).

**Figure 1:**
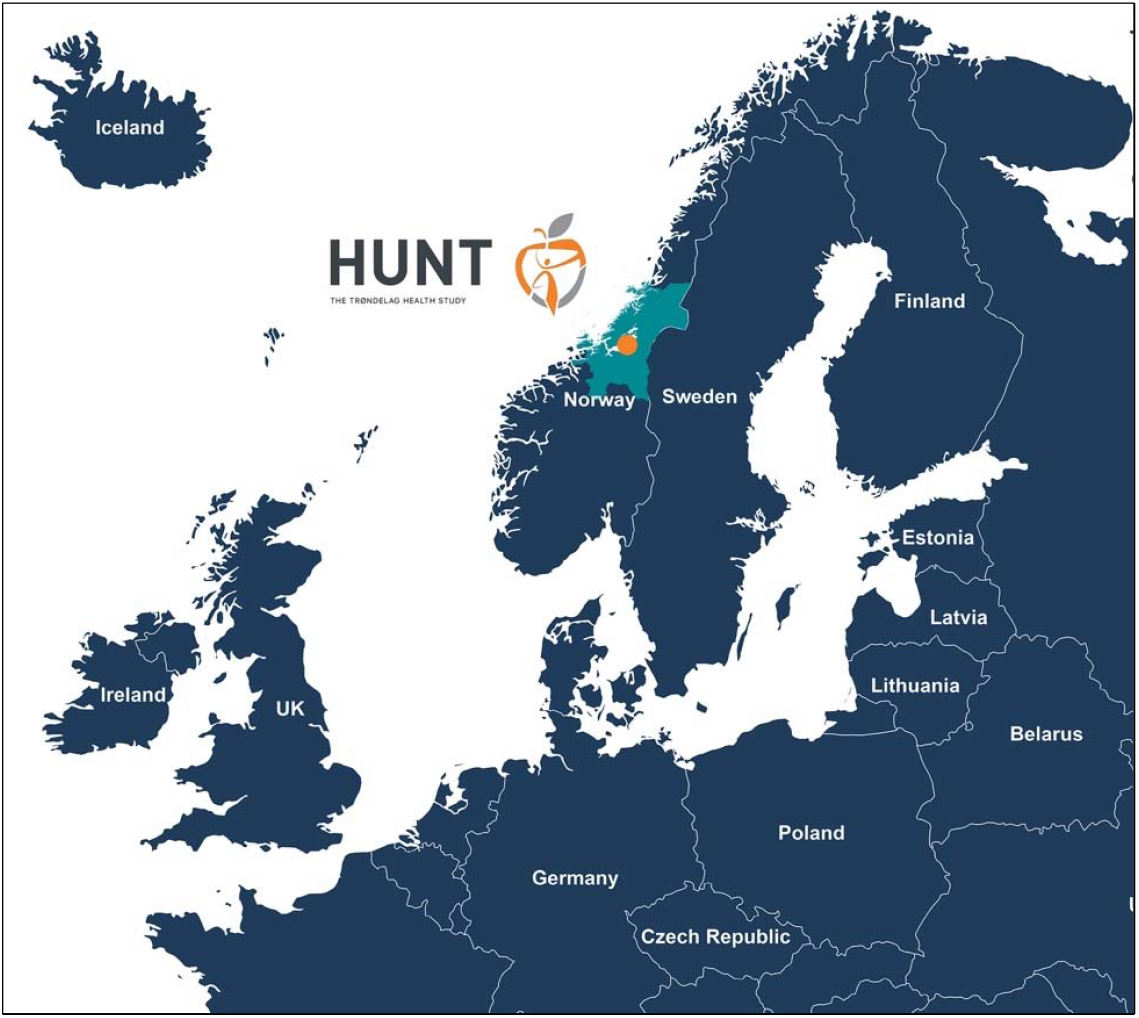
The Trøndelag Health Study (HUNT), Trøndelag, Norway. The country of Trøndelag is shaded light blue and the orange point indicates the location of the HUNT Research Center at Levanger.

To understand the genetic basis of diseases, as well as follow individuals with genetic and epidemiological risk factors in a well-ascertained county in Norway, we established a comprehensive collaboration in 2005 between the HUNT study at the Norwegian University of Science and Technology, Norway and the University of Michigan, USA. This paper presents the history and status of this collaboration by describing the study population, the strategy incorporating genotyping, sequencing, and imputation-based approaches in HUNT, the vast phenotype data collected by decades of HUNT researchers, the linkage to the digitized public health care system and key findings to date.

### Study population

HUNT is an ongoing population-based health study in Trøndelag County, Norway. The study collects health-related data from questionnaires, interviews and clinical examinations from individuals within this geographical region (**Figure 2**). More than 229 000 adults (20 years or older at recruitment) have participated in the study to date, of whom 95 000 have provided at least one biological sample (https://www.ntnu.edu/hunt/hunt-samples)^1-4^. The periodic survey design includes four recruitment waves. HUNT1 (1984-86), HUNT2 (1995-97), HUNT3 (2006-08) and HUNT4 (2017-19) concentrated primarily on the North-Trøndelag area, where all adults (age ≥20 years) were invited. In addition, HUNT4 expanded to collect basic questionnaire data from the adult population of South-Trøndelag (105,797 additional participants)^3^. ∼19 000 adults have participated in all four HUNT waves, thus having longitudinal questionnaire and physical exam information spanning over 35 years. Complementing the surveys in adult participants, four separate Young-HUNT surveys gathered data from ∼25 000 adolescents in junior high and high school, concurrent with HUNT2-4. No genotyping has been performed on Young-HUNT, however 4 212 have sequentially participated in the adult version of HUNT. The HUNT Study has a high level of participation (ranging from 54% to 89% between surveys among those invited) making the cohort a good representative of the general Norwegian population. The HUNT and Young-HUNT cohorts are described in more detail elsewhere^1-5^.

**Figure 2:**
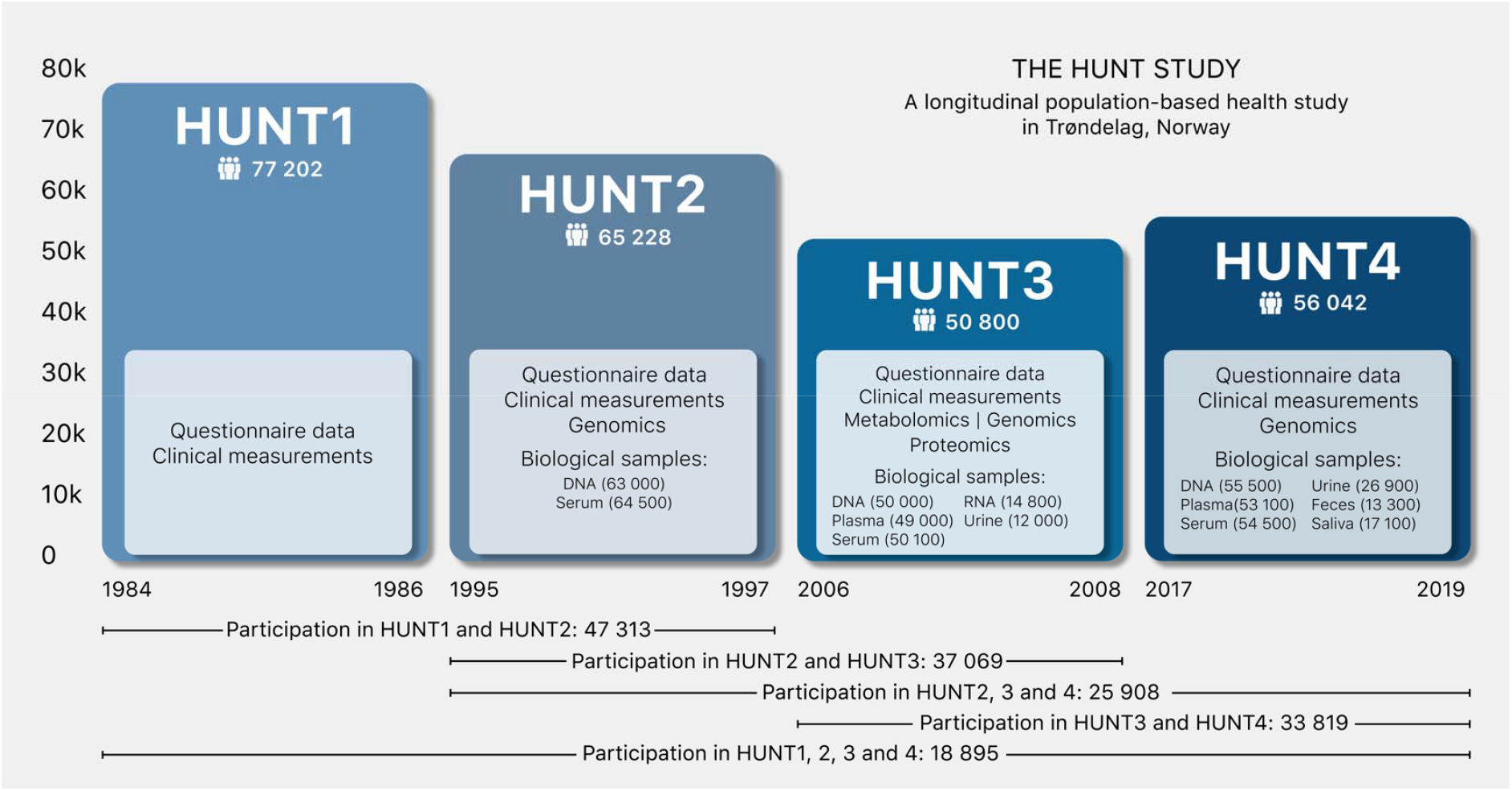
Sample sizes across the HUNT1-4 surveys and details of key data and biological samples. DNA: Deoxyribonucleic acid, HUNT: Trøndelag Health Study, RNA: Ribonucleic acid

### Genotyping and imputation study design in HUNT

∼88 000 individuals provided DNA for medical research during at least one of the HUNT recruitment periods. Initially, our efforts were focused on identifying genetic variants associated with myocardial infarction (MI)^6-8^. Towards this goal, we genotyped exome variants and performed low-pass whole-genome sequencing (4.7x average coverage) in 2014 on 2,201 samples from HUNT2 and HUNT3 (HUNT-WGS) (**Supplementary Table 1**), including early-onset MI cases and equal numbers of sex- and age-matched controls. Although no novel significant associations were found, likely due to the limited sample size, this set of low-pass sequences provided important insights into genetic variants present in the Norwegian population and contributed Norwegian reference sequences to the Haplotype Reference Consortium (HRC) imputation panel^9^. We next undertook genome-wide genotyping on all HUNT2-3 participants (n=71 860) with available DNA (**Figure 3)**. Motivated by a goal of capturing high-quality, common- and low-frequency and Norwegian-specific variants, we used a variety of approaches to observe or estimate genotypes: 1) direct genotyping using standard and customized HumanCoreExome arrays from Illumina; 2) genotyping and imputation with a merged HRC and HUNT-WGS imputation panel; and 3) imputation with the TOPMed imputation panel (**Figure 4**). After genotyping 12 864 with standard HumanCoreExome arrays (HumanCoreExome 12 v1.0 and v1.1), we performed genotyping on the remaining samples using a customized HumanCoreExome array (UM HUNT Biobank v1.0) which included protein-altering variants observed in HUNT-WGS. This resulted in genotyping 358 964 polymorphic variants. We next used the 2 201 sequenced samples (HUNT-WGS) for joint imputation with the HRC panel^10^. We previously showed that imputation with a HUNT-specific reference panel improved imputation of low-frequency and population-specific variants compared to either using the 1000 Genomes or HRC reference panels alone^11^. Lastly, we imputed 25 million variants from the TOPMed imputation panel (minor allele count greater than 10), which resulted in slightly lower imputation quality compared to the population-specific reference panel but captured a larger number of variants (**Supplementary Figure 1**). These two imputed datasets can be used separately in downstream analysis; we recommend using the HRC and HUNT-WGS imputation for the investigation of the Norwegian specific variants. Together, the imputations resulted in 33 million variants in 70 517 individuals from HUNT2 or HUNT3 of which 3.3 million variants are not found in UK Biobank. Finally, 18 722 new samples from HUNT4 have recently been genotyped using the same approaches (HumanCoreExome array, UM HUNT Biobank v2.0) and following imputation will create a new, larger data freeze of ∼88,000 individuals from HUNT2-4. Further details of the quality control and imputation in HUNT can be found in the **Supplementary Note**.

**Figure 3:**
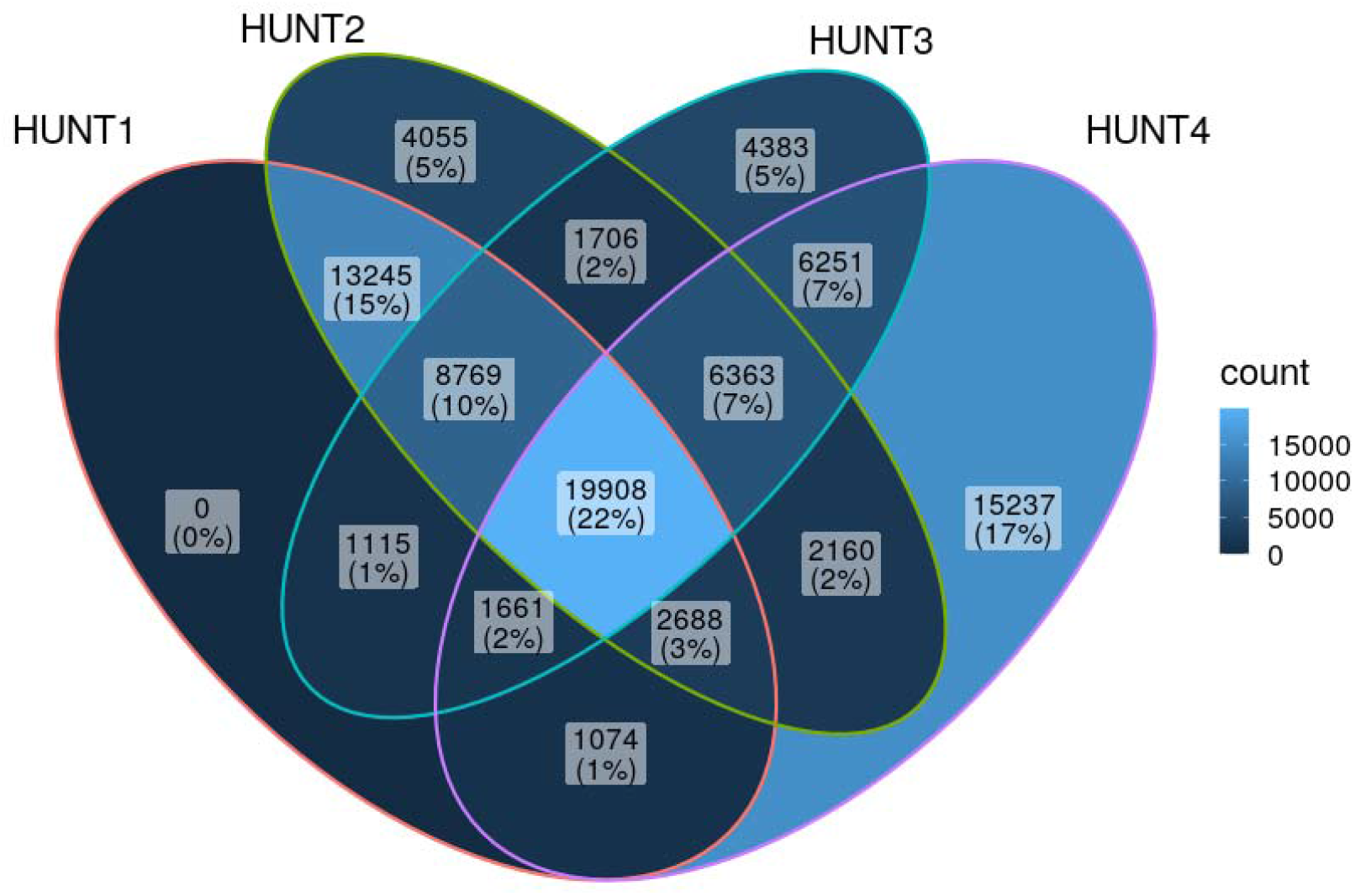
Genotyped samples from HUNT available from the different HUNT surveys (n=88 615). HUNT: Trøndelag Health Study

**Figure 4:**
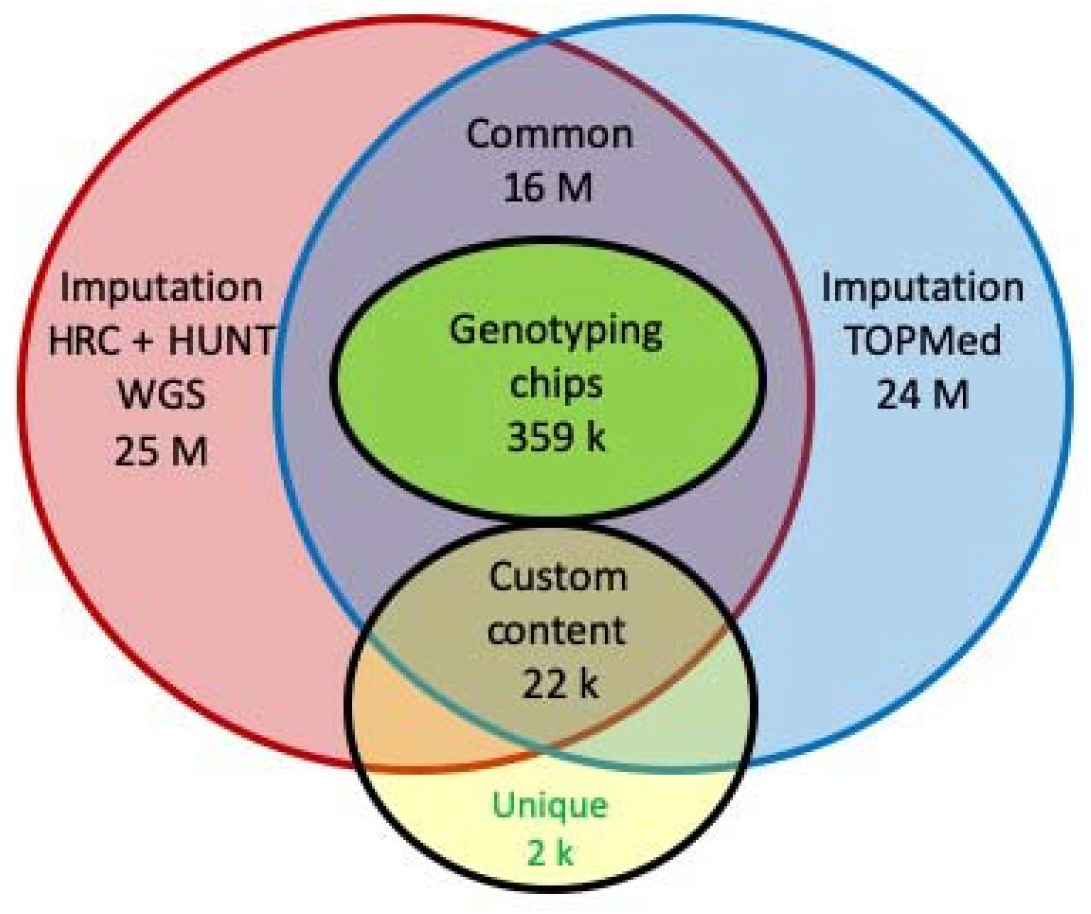
Genotyping and imputation-based approach in HUNT. HUNT: Trøndelag Health Study, HRC: Haplotype Reference Consortium, k: Thousand, M: Million, TOPMed: Trans-Omics for Precision Medicine, WGS: Whole-genome sequencing

### Phenotypes

A broad range of phenotypes are available for HUNT participants based on laboratory tests, clinical examinations, and self-reported questionnaires. These include non-fasting blood lipids and glycaemic traits; history (including age of diagnosis) of a range of diseases including cardiovascular events; basic demographics including sex and participation age; anthropometrics including weight, height, BMI, and waist-to-hip ratio; blood pressure measurements; and lifestyle information including smoking status (**Table 1**). HUNT data categories have been previously described^2, 3^, and are described in detail on the HUNT databank website (https://www.ntnu.edu/hunt/databank). Importantly, many measurements and questionnaire items have been intentionally kept identical or similar across HUNT surveys to enable longitudinal analyses.

**Table 1:**
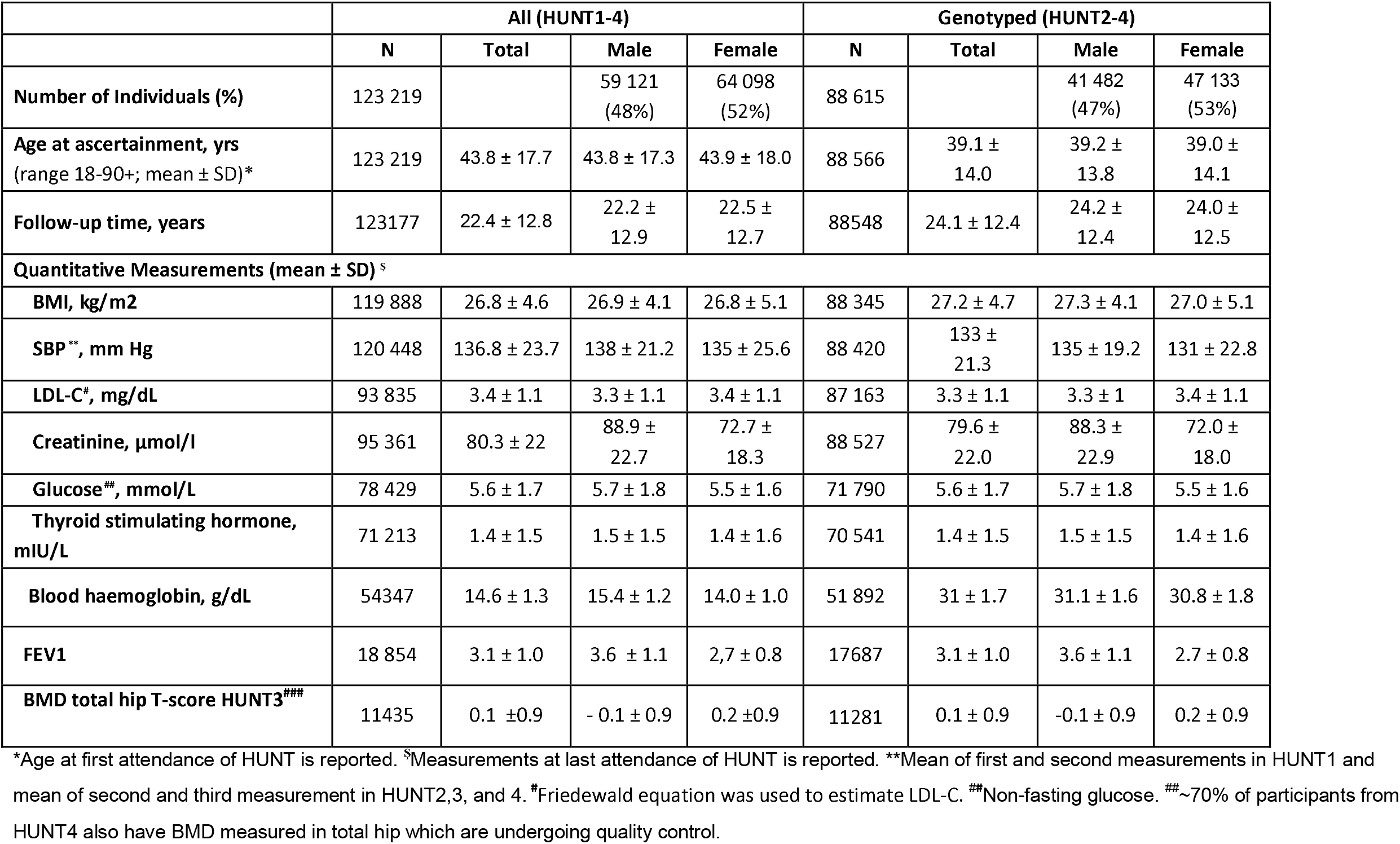
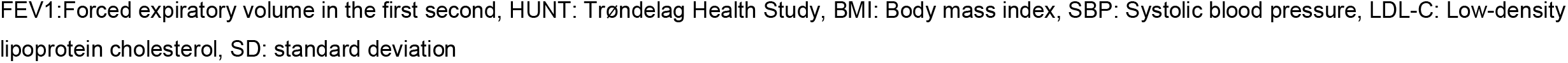
HUNT cohort demographics for all attendees at HUNT1-4 clinical examinations (n=123 219), and among those genotyped (n=88 615).

|  | All (HUNT1-4) |  |  |  | Genotyped (HUNT2-4) |  |  |  |
| --- | --- | --- | --- | --- | --- | --- | --- | --- |
|  | N | Total | Male | Female | N | Total | Male | Female |
| **Number of Individuals (%)** | 123 219 |  | 59 121<br>(48%) | 64 098<br>(52%) | 88 615 |  | 41 482<br>(47%) | 47 133<br>(53%) |
| **Age at ascertainment, yrs**<br>(range 18-90+; mean ± SD)* | 123 219 | 43.8 ± 17.7 | 43.8 ± 17.3 | 43.9 ± 18.0 | 88 566 | 39.1 ±<br>14.0 | 39.2 ±<br>13.8 | 39.0 ±<br>14.1 |
| **Follow-up time, years** | 123177 | 22.4 ± 12.8 | 22.2 ±<br>12.9 | 22.5 ±<br>12.7 | 88548 | 24.1 ± 12.4 | 24.2 ±<br>12.4 | 24.0 ±<br>12.5 |
| **Quantitative Measurements (mean ± SD) <sup>§</sup>** |  |  |  |  |  |  |  |  |
| **BMI, kg/m<sup>2</sup>** | 119 888 | 26.8 ± 4.6 | 26.9 ± 4.1 | 26.8 ± 5.1 | 88 345 | 27.2 ± 4.7 | 27.3 ± 4.1 | 27.0 ± 5.1 |
| **SBP \*\*, mm Hg** | 120 448 | 136.8 ± 23.7 | 138 ± 21.2 | 135 ± 25.6 | 88 420 | 133 ±<br>21.3 | 135 ± 19.2 | 131 ± 22.8 |
| **LDL-C #, mg/dL** | 93 835 | 3.4 ± 1.1 | 3.3 ± 1.1 | 3.4 ± 1.1 | 87 163 | 3.3 ± 1.1 | 3.3 ± 1 | 3.4 ± 1.1 |
| **Creatinine, μmol/l** | 95 361 | 80.3 ± 22 | 88.9 ±<br>22.7 | 72.7 ±<br>18.3 | 88 527 | 79.6 ±<br>22.0 | 88.3 ±<br>22.9 | 72.0 ±<br>18.0 |
| **Glucose ##, mmol/L** | 78 429 | 5.6 ± 1.7 | 5.7 ± 1.8 | 5.5 ± 1.6 | 71 790 | 5.6 ± 1.7 | 5.7 ± 1.8 | 5.5 ± 1.6 |
| **Thyroid stimulating hormone, mIU/L** | 71 213 | 1.4 ± 1.5 | 1.5 ± 1.5 | 1.4 ± 1.6 | 70 541 | 1.4 ± 1.5 | 1.5 ± 1.5 | 1.4 ± 1.6 |
| **Blood haemoglobin, g/dL** | 54347 | 14.6 ± 1.3 | 15.4 ± 1.2 | 14.0 ± 1.0 | 51 892 | 31 ± 1.7 | 31.1 ± 1.6 | 30.8 ± 1.8 |
| **FEV1** | 18 854 | 3.1 ± 1.0 | 3.6 ± 1.1 | 2.7 ± 0.8 | 17687 | 3.1 ± 1.0 | 3.6 ± 1.1 | 2.7 ± 0.8 |
| **BMD total hip T-score HUNT3 ###** | 11435 | 0.1 ± 0.9 | - 0.1 ± 0.9 | 0.2 ± 0.9 | 11281 | 0.1 ± 0.9 | -0.1 ± 0.9 | 0.2 ± 0.9 |

\*Age at first attendance of HUNT is reported. <sup>§</sup>Measurements at last attendance of HUNT is reported. \*\*Mean of first and second measurements in HUNT1 and mean of second and third measurement in HUNT2,3, and 4. #Friedewald equation was used to estimate LDL-C. ##Non-fasting glucose. ###~70% of participants from HUNT4 also have BMD measured in total hip which are undergoing quality control.
FEV1: Forced expiratory volume in the first second, HUNT: Trøndelag Health Study, BMI: Body mass index, SBP: Systolic blood pressure, LDL-C: Low-density lipoprotein cholesterol, SD: standard deviation

### Linkage to regional and national health registries

Using the unique personal identification number given to all Norwegian citizens allows for longitudinal follow-up by linkage between HUNT data, regional and national registries and electronic health records. Norway currently has 17 national health registries (https://helsedata.no/no/) that are mandatory and cover the entire population (**Supplementary Table 2**). Commonly used national registries linked with HUNT include the Norwegian Cause of Death Registry (established 1951), the Cancer Registry of Norway (established 1952), the Medical Birth Registry of Norway (established 1967), and the Norwegian Prescription Database (established 2004). Another 52 national disease-specific medical quality registries hold detailed information on treatment and responses at an individual level (https://www.kvalitetsregistre.no/registeroversikt) (**Supplementary Table 2**). Electronic health records from the local hospitals hold International Statistical Classification of Diseases and Related Health Problems (ICD) codes back to 1987. Potential linkage to non-health related registries expands the data resource, that among others includes Statistics Norway, recording income and wealth statistics for individuals and households, and the Norwegian Armed Forces Health Registry (https://helsedata.no/no/). Together, the listed registries provide opportunities to integrate a breadth of data from multiple time points to obtain high quality phenotypes and related information on, for example, environmental and socioeconomic factors. Time-stamped data allows studies of disease development and progression. Some selected disease endpoints are presented in **Table 2**.

**Table 2:** ICD codes captured in the local hospital register from 1987 to 2021 for selected diseases and the observed case numbers in genotyped HUNT participants.*

| ICD Chapter | ICD-9 | ICD-10 | Cases genotyped |
| --- | --- | --- | --- |
| **Infectious and parasitic diseases** |  |  |  |
| COVID-19, virus identified |  | U07.1 | 39 |
| Personal history of COVID-19 |  | U08 | 112 |
| Post COVID-19 condition |  | U09 | 8 |
| **Neoplasms** |  |  |  |
| Malignant neoplasm of colon, rectosigmoid junction, rectum, anus and anal canal | 153, 154 | C18, C19, C20, C21 | 3291 |
| Malignant neoplasm of bronchus and lung | 162 | C34 | 2521 |
| Malignant melanoma of skin | 172 | C43 | 2345 |
| Malignant neoplasm of breast | 174, 175 | C50 | 2047 |
| Malignant neoplasm of prostate | 185 | C61 | 3153 |
| **Endocrine, nutritional and metabolic diseases** |  |  |  |
| Hypothyroidism | 240, 241, 242, 243, 244, 245 | E00, E01, E02, E03 | 7250 |
| Type 2 diabetes mellitus | 250 | E11 | 11444 |
| **Mental and behavioural disorders** |  |  |  |
| Dementia | 290, 294, 331 | F00, F01,<br>F02, F03,<br>G30, G31.1 | 4509 |
| Mood (affective) disorders | 296, 298, 300, 301,<br>311 | F30, F31,<br>F32, F33,<br>F34, F38,<br>F39 | 9267 |
| **Diseases of the nervous system** |  |  |  |
| Parkinson's disease | 332, 333 | G20, G21,<br>G22, F02.3 | 5124 |
| Epilepsy | 345 | G40 | 11480 |
| Migraine | 346 | G43 | 1728 |
| **Diseases of the eye and adnexa** |  |  |  |
| Glaucoma | 365 | H40 | 4223 |
| **Diseases of the circulatory system** |  |  |  |
| Essential (primary) hypertension | 401 | I10 | 18263 |
| Angina pectoris | 413.9 | I20 | 8388 |
| Acute myocardial infarction | 410 | I21 | 10144 |
| Atrial fibrillation and flutter | 427 | I48 | 14538 |
| Heart failure | 428 | I50, I09,<br>I11 | 7273 |
| Intracerebral hemorrhage | 431, 432 | I61 | 2901 |
| Aortic aneurysm and dissection | 441 | I71 | 2491 |
| **Diseases of the respiratory system** |  |  |  |
| Chronic obstructive pulmonary disease | 496 | J44.8,<br>J44.9 | 6529 |
| Asthma | 493 | J45 | 6511 |
| Post-inflammatory pulmonary fibrosis | 515 | J84.1,<br>J84.8 | 954 |
| **Diseases of the digestive system** |  |  |  |
| Crohn's disease | 555 | K50 | 586 |
| Ulcerative colitis | 556 | K51 | 2288 |
| Coeliac disease | 579 | K90.0 | 1152 |
| **Diseases of the skin and subcutaneous tissue** |  |  |  |
| Atopic dermatitis | 691.8 | L20 | 1067 |
| Psoriasis | 696 | L40 | 3135 |
| **Diseases of the musculoskeletal system and connective tissue** |  |  |  |
| Gout | 274 | M10 | 6178 |
| Ankylosing spondylitis | 720 | M45 | 2064 |
| **Diseases of the genitourinary system** |  |  |  |
| Chronic kidney disease | 585 | N18 | 4668 |
| **Pregnancy, childbirth and the puerperium** |  |  |  |
| Gestational hypertension | 342 | O13 | 1666 |
| **Prescription data <sup>#</sup>** |  |  |  |
| Low dose aspirin | - | - | 22500 |
| Statin | - | - | 2200 |

\*Numbers are from a data query (August 8, 2021) from the Nord-Trøndelag Hospital Trust including Namsos and Levanger Hospitals of participants selected for genotyping from HUNT2-4. The register is ongoing and therefore the number of cases continue to increase over time.
<sup>#</sup>Approximate numbers from the prescription register and restricted to those genotyped in HUNT2-3 only.
HUNT: Trøndelag Health Study, ICD: International Statistical Classification of Diseases and Related Health Problems

### Analytical approaches with related samples

97% of HUNT participants are of Norwegian ancestry^4^. Using principal components of ancestry projected onto the Human Genome Diversity Project, we typically exclude samples of non-European ancestry (<2%) (**Supplementary Figure 2**) due to limited power. We have observed fine-scale differences between North- and South-Trøndelag and between individuals born closer to the coast versus the border with Sweden^12^. Additionally, because of high ascertainment from a single county in Norway (Trøndelag), there are many related individuals within the cohort. 79 551 (89%) out of 88 615 HUNT2-4 participants have at least one 2nd degree or closer relative who also participates in HUNT (**Supplementary Figure 3, Supplementary Table 3**). High degree of participant relatedness in the dataset on one hand allows for unique data-analysis methods using nuclear or extended families, but can result in bias when using methods that assume unrelated individuals or power loss if related individuals are exclude. An early effort to use extended families and genetic data in HUNT was for the analysis of rare coding variants^13^, where family samples can provide more power to detect associations when sample sizes were limited and only a modest fraction of all trait-associated variants were identified^13^.

Previously, methods had been developed to account for relatedness for analysis of quantitative traits^14^, but methods to properly account for relatedness and control for unbalanced case-control ratios for binary traits were lacking. We therefore developed statistical methods to allow for the analysis of all individuals, and to control for case-control imbalance of binary phenotypes, which is commonly observed in biobanks such as HUNT. These methods, which are computationally efficient in biobank-scale data, allowed us to perform association testing in HUNT for both single variants (using SAIGE) and gene-based burden tests (using SAIGE-GENE), while accounting for sample relatedness with a sparse identical by state (IBS) sharing matrix^13, 15-17^. These methods account for case-control imbalance of binary phenotypes, typical in a population-based sample, by using the saddlepoint approximation to calibrate unbalanced case-control ratios in score tests based on logistic mixed models [Add SAIGE reference]. We demonstrated a vast improvement in reducing type I error rates when analyzing unbalanced case control ratios with SAIGE in HUNT. For example, venous thromboembolism with 2 325 cases and 65 294 controls and a case:control ratio of 0.036 had substantial inflation of type I error with methods available prior to the development of SAIGE (**Supplementary Figure 4**). To demonstrate the application of SAIGE-GENE, we investigated 13 416 genes, with at least two rare (MAF□≤□1%) missense and/or stop-gain variants that were directly genotyped or imputed from the joint HRC and HUNT-WGS reference panel among 69 716 Norwegian samples from HUNT2-3 with measured high-density lipoprotein. We identified eight genes with p-values below the exome-wide significance threshold (P□≤□2.5□×□10^−6^), seven of which remained significant after conditioning on nearby single-variant associations, suggesting independent rare coding variants within these genes^16^. Importantly, using SAIGE and SAIGE-GENE, we were able to use all samples, account for sample relatedness case-control imbalance, and maintain well controlled type I error rates.

A traditional way of using related samples is linkage analysis, which however has computational challenges in the era of whole-genome genetics. To allow for linkage testing in datasets with millions of genetic markers, faster and computationally scalable linkage analysis method have been developed, Population Linkage^18^. Population Linkage uses a Haseman-Elston regression (originally used for sibling pair linkage analysis) to estimate variance components from pairwise relationships and IBD estimates. Using HUNT data, Zajac et. al. observed 25 significant linkage peaks with LOD > 3 across 19 distinct loci for the four traits (high-density lipoprotein, low-density lipoprotein, total cholesterol and triglycerides), where 5 peaks with LOD > 3 were not replicated at genome-wide significance in a genome-wide association study (GWAS) of 359,432 genotyped variants in HUNT. However, after imputing the dataset with the HRC and HUNT-WGS reference panel to cover more variants or meta-analysis in GLGC, significant associations in all 5 linkage peaks were observed. This study demonstrates one of the benefits of linkage analysis over GWAS, that is the ability to test for linkage in regions that are difficult to genotype such as rare variants, structural variants, copy number variants or variants in highly repetitive regions, as long as identical by descent segments in the region can be identified^18^. Finally, linkage analysis may improve statistical power when investigating rare risk variants which segregate within families and reduce confounding effects of population stratification.

The high degree of relatedness in the HUNT Study participants has enabled analysis methods are tailored to this study design. These include GWAS by proxy^19, 20^, in which the phenotypes of non-genotyped family members of genotyped HUNT participants can be used to identify proxy-cases, individuals with a proportion (0.5 for first degree relatives) of the genetic risk of cases. These proxy-cases can be appropriately modelled to increase statistical power in GWAS. For example, the power to detect an allele with an odds ratio of 1.1 and MAF of 0.21 at an alpha of 5×10^−8^ increases from 0.419 to 0.644 when proxy-cases are appropriately modelled instead of used in controls as in standard GWAS (**Supplementary Figure 5**).

### Genetic discoveries from HUNT

The wealth of phenotypic and genetic data available in the HUNT cohort has led to the discovery of many new genetic associations across a broad range of traits (**Table 3**). Early genetic studies of HUNT participants used exome arrays and focused on cardiovascular disease. We identified a novel coding variant in *TM6SF2* associated with total cholesterol, MI, and liver enzymes^6^ and replicated known MI associations at the 9p21 locus and a low-frequency missense variant in the *LPA* gene (p.Ile1891Met)^7^. Following the genotyping of nearly 70 000 participants in HUNT2 and HUNT3 and the development of a combined HRC and HUNT-WGS imputation reference panel, we extended our analyses to a genome-wide search (**Figure 5**). Through imputation of indels called from low-pass HUNT-WGS, we discovered a rare mutation in the *MEPE* gene, enriched in the Norwegian population (0.8% in HUNT, 0.1% in non-Finnish Europeans), that was associated with low forearm bone mineral density and increased risk of osteoporosis and fractures^21^. Although this region had been previously identified as associated with bone mineral density^22^, the association in HUNT with replication in the UK biobank^23^ pin-pointed *MEPE* as the likely causal gene in the region by identifying an insertion/deletion polymorphism that likely resulted in a loss-of-function protein. In another study we paid special attention to loss-of-function mutations associated with favorable blood lipid profiles (reduced LDL cholesterol and reduced CAD risk) which were not associated with altered liver enzymes or liver damage. We additionally found one elderly individual with homozygous *ZNF529* loss-of-function variant showing no signs of cardiovascular disease or diabetes, suggesting that the full knock-out of this gene is viable. This highlighted *ZNF529* as a potential therapeutic target for lipids^24^ identified from sequencing and custom content genotyping.

**Table 3:** Genetic discoveries across HUNT genotyping and analysis strategies.

| Strategy | Frequency range/<br>Number of<br>variants | Benefits | Exemplary<br>papers |
| --- | --- | --- | --- |
| Genotyping with custom exome content designed from HUNT sequenced samples (UM HUNT Biobank Array) | Rare-Common/<br>80,137-358,964 | Identify low-frequency variants not amenable to imputation-based approaches | Identified likely causal gene, *TM6SF2*, associated with TC and MI<sup>33</sup> |
|  |  |  | Found LOF variant in *ZNF529* that leads to lower LDL-C<sup>24</sup> |
| HRC and HUNT-WGS Imputation from Human CoreExome Array | Low-Common/<br>22 million | Include population-specific variants through improved imputation | Population-enriched variant in *MEPE* pinpoints causal gene for fracture risk<sup>21</sup> |
|  |  |  | Identified variants associated with thyroid function<sup>29</sup>, kidney function<sup>27</sup> serum PCSK9<sup>34</sup> and atrial fibrillation<sup>25, 26</sup> |
| TOPMed Imputation from Human CoreExome Array | Low-Common/<br>25 million | Expand number of available variants for association testing | Identified variants associated with troponin and serum iron in the general population<sup>31, 32</sup> |
| Family-based design, > 15,000 sibling pairs, >35,000 parent-offspring | Any/<br>10-1000 | Improve effect size estimates and test traits among un-studied relatives | Introduced new analysis methods, including SAIGE<sup>35</sup>, within-family Mendelian randomization and GWAS<sup>36, 37</sup> |
| Mendelian Randomization | Any/<br>10-1000 | Identify causal links between environmental factors (genetically determined traits) and outcomes | Explored the role of lipids and apolipoproteins on kidney-function<sup>38, 39</sup> |
|  |  |  | Demonstrated an inverse association between thyroid stimulating hormone and thyroid cancer<sup>29</sup> |

Note: Rare variants <1% minor allele frequency (MAF), Low frequency variants 1-5% MAF, Common variants >5% MAF
HUNT: Trøndelag Health Study, HRC: Haplotype Reference Consortium, LDL-C: Low density lipoprotein cholesterol, LOF: Loss-of-function, MI: Myocardial infraction, TC: Total cholesterol, TOPMed: Trans-Omics for Precision Medicine, UM: University of Michigan, WGS: Whole-genome sequencing

**Figure 5:**
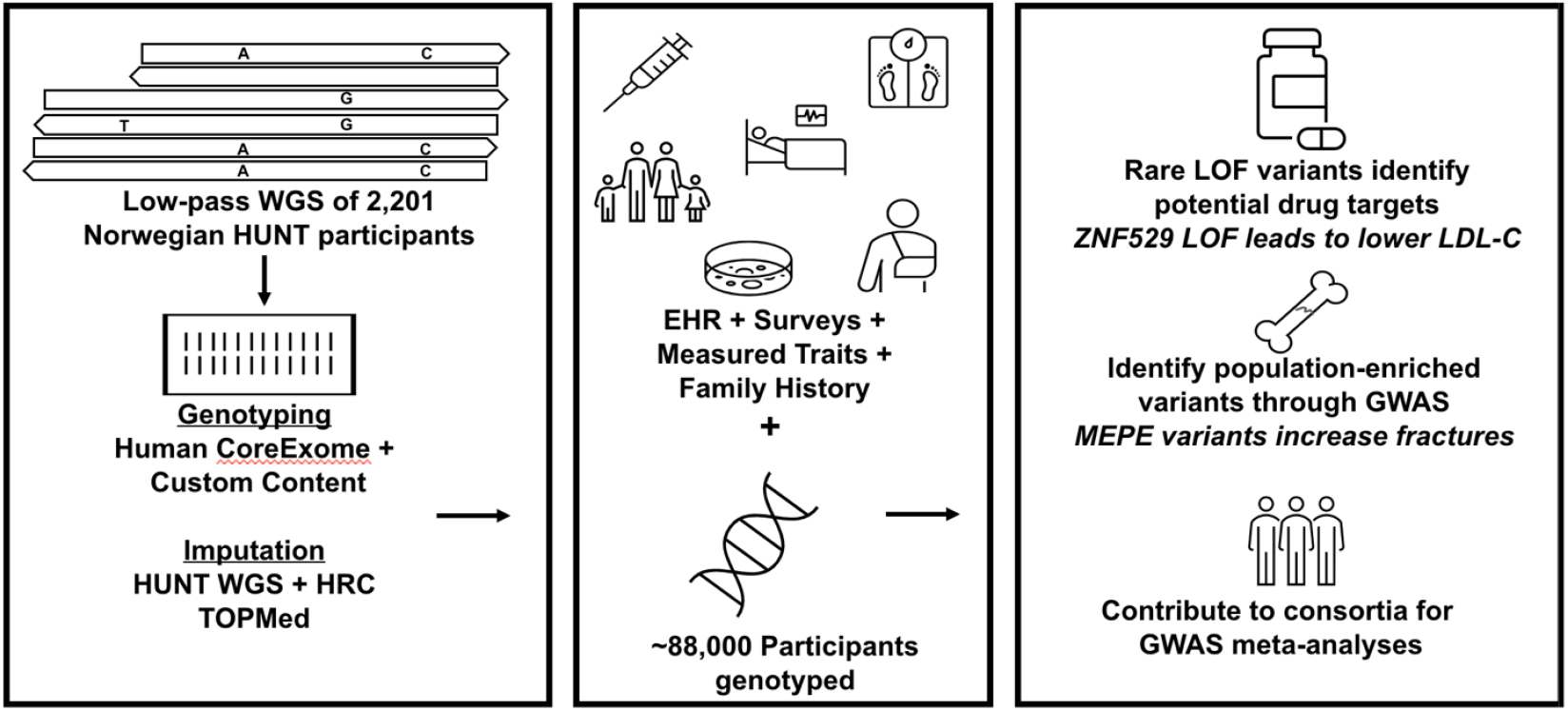
Overview of HUNT genetics, phenotypic resources and discoveries. EHR: Electronic health records, GWAS: Genome-wide association study, HUNT: Trøndelag Health Study, WGS: Whole genome sequencing, HRC: Haplotype Reference Consortium, LDL-C: Low density lipoprotein cholesterol, LOF: Loss-of-function, TOPMed: Trans-Omics for Precision Medicine

On top of the association studies performed using HUNT data only, we have contributed to many international consortium efforts aimed at aggregating GWAS data across cohorts. By performing GWAS meta-analyses that included HUNT and other cohorts, efforts driven by our research team have identified genetic variants associated with atrial fibrillation which may act through a mechanism of impaired muscle cell differentiation and tissue formation during fetal heart development^25^ and cardiac structural remodeling^26^; variants associated with estimated glomerular filtration rate exhibiting a sex-specific effect^27, 28^; and variants associated with thyroid stimulating hormone that revealed an inverse relationship between TSH levels and thyroid cancer^29^. Later studies using the TOPMed reference panel^30^ identified variants associated with circulating cardiac troponin I level and investigated its role as a non-causal biomarker for MI using Mendelian randomization^31^, and identified variants associated with iron-related biomarker levels and explored their relationship with all-cause mortality^32^.

### Causal inference and family effects

The high degree of relatedness in the HUNT Study offers a unique opportunity to use family-based designs to investigate causal associations. Mendelian randomization (MR), which uses genetic variants as instrumental variables to investigate modifiable (non-genetic) factors, was first proposed using parent-offspring^40^. Alleles that are inherited from each parent are randomly determined during the meiotic process. This random allocation is essential to providing reliable comparisons in MR studies. However, due to the lack of genotyped family data, previous studies applied MR on the population-level, where the random allocation of alleles is only approximate. We were able to use the ∼15 000 families in HUNT to perform MR as originally proposed - in family-based designs^36^. Using this approach in HUNT, we showed empirically that MR estimates from samples of unrelated individuals for the association of taller height and lower BMI increase educational attainment, were likely induced by population structure, assortative mating or dynastic effects. We observed no clear associations in within-family MR analyses in HUNT or in a replication cohort of 222 368 siblings from 23andMe^36^. This approach has since grown in popularity and, together with HUNT, many cohorts now contribute to the investigation of causal associations with family-based designs^37^.

Further leveraging the family structure information in HUNT, we have performed and have future opportunities to investigate causal effects between family members, for example parent-offspring effects^41, 42^, assortative mating and sibling effects^43^. These study designs have not been previously possible due to the lack of genotyped family data and this has both limited causal inference (as mentioned above) and the ability of typical GWAS to distinguish between direct and indirect genetic effects^37^. HUNT data allows for study designs to disentangle these sources of genotype-phenotype associations in humans. In one such example, we used 26 057 mother–offspring and 9 792 father–offspring pairs to investigate whether adverse environmental factors *in utero* increased future risk of cardiometabolic disease in the offspring. We observed that adverse maternal intrauterine environment, as proxied by maternal SNPs that influence offspring birthweight, were unlikely to be a major determinant of late-life cardiometabolic outcomes of the offspring^41^.

## Data Availability

Researchers associated with Norwegian research institutes can apply for the use of HUNT data and samples with approval by the Regional Committee for Medical and Health Research Ethics. Researchers from other countries may apply if collaborating with a Norwegian Principal Investigator. Information for data access can be found at https://www.ntnu.edu/hunt/data. The HUNT variables are available for browsing on the HUNT databank at https://hunt-db.medisin.ntnu.no/hunt-db/. Data linkages between HUNT and health registries require that the principal investigator has obtained project-specific approval for such linkage from the Regional Committee for Medical and Health Research Ethics, Norway and each registry owner.

https://hunt-db.medisin.ntnu.no/hunt-db

## Contribution to collaborative studies

We contribute to genetic studies worldwide through participation in consortia focused on a variety of diseases including cardiovascular disease^44, 45^, lipids^46, 47^, type 2 diabetes^48^, osteoporosis^49^, decline in kidney function^50^, Alzheimer’s disease^51^, bipolar disease^52^, intracranial aneurysms^53^, insomnia^54^, respiratory health^55^ and sleepiness^56^. We also contributed HUNT data to studies of anthropometric traits^57^, alcohol and nicotine use^58, 59^, COVID-19^60^, phenome-wide discovery^61^, and genetic risk prediction^62^, among others. We believe that team science by consortia^61^ fulfills the goals of the HUNT study and moves the science fastest towards new discoveries and improved human health.

## Summary

Together, the multifaceted genetic discovery strategy incorporating genotyping, sequencing, and imputation-based approaches in HUNT has aided the identification of likely causal genes and variants for disease and human traits. It has also proved to be a valuable resource for genetically informed methods of causal inference, supporting the identification of modifiable risk factors. We owe this success to the willingness and high participation rates of the people of Trøndelag, the vast phenotyping collected by decades of HUNT researchers, and access to digitized public health care systems. We anticipate that the rich data collection will continue to be a unique dataset for future opportunities in longitudinal and family-based designs, genetic discoveries, Mendelian randomization, meta-analysis and polygenic score validation, well into the future.

## HUNT Data Access

Researchers associated with Norwegian research institutes can apply for the use of HUNT data and samples with approval by the Regional Committee for Medical and Health Research Ethics. Researchers from other countries may apply if collaborating with a Norwegian Principal Investigator. Information for data access can be found at https://www.ntnu.edu/hunt/data. The HUNT variables are available for browsing on the HUNT databank at https://hunt-db.medisin.ntnu.no/hunt-db/. Use of the full genetic data set requires the use of an approved secure computing solution such as the HUNT Cloud (https://docs.hdc.ntnu.no/). Data linkages between HUNT and health registries require that the principal investigator has obtained project-specific approval for such linkage from the Regional Committee for Medical and Health Research Ethics, Norway and each registry owner. GWAS summary statistics from publications including HUNT are available from NTNU Open Research Data (https://dataverse.no/dataverse/root) and the Willer lab (http://csg.sph.umich.edu/willer/public/).

## Ethics

The genotyping in HUNT and work presented in this cohort profile was approved by the Regional Committee for Ethics in Medical Research, Central Norway (2014/144, 2018/1622, 152023). All participants signed informed consent for participation and the use of data in research.

## Acknowledgement

A special thanks to all HUNT participants for donating their time, samples and information to help others.

The Trøndelag Health Study (HUNT) is a collaboration between HUNT Research Centre (Faculty of Medicine and Health Sciences, NTNU, Norwegian University of Science and Technology), Trøndelag County Council, Central Norway Regional Health Authority, and the Norwegian Institute of Public Health. The genotyping in HUNT was financed by the National Institutes of Health; University of Michigan; the Research Council of Norway; the Liaison Committee for Education, Research and Innovation in Central Norway; and the Joint Research Committee between St Olavs hospital and the Faculty of Medicine and Health Sciences, NTNU. The genetic investigations of the HUNT Study is a collaboration between researchers from the K.G. Jebsen Center for Genetic Epidemiology, NTNU and the University of Michigan Medical School and the University of Michigan School of Public Health. The K.G. Jebsen Center for Genetic Epidemiology is financed by Stiftelsen Kristian Gerhard Jebsen; Faculty of Medicine and Health Sciences, NTNU, Norway.

We want to thank clinicians and other employees at Nord-Trøndelag Hospital Trust for their support and for contributing to data collection in this research project.

Thanks to Trøndelag County Council, Jon Olav Sliper for creating Figure 1, K.G. Jebsen Center Communications Officer Janne Tellefsen for creating Figure 2 and Willer Lab Research Coordinator Bethany Klunder for organizing our collaborative meetings.

The authors thank Robin Walters and Mark Daly for an internal review of this manuscript prior to submission.

We also acknowledge; **HUNT-MI Leadership:** Kristian Hveem, Cristen J. Willer, Oddgeir L. Holmen, Michael Boehnke, Gonçalo R. Abecasis, Bjorn Olav Åsvold, Ben M. Brumpton; **Scientific Advisory Committee:** Ele Zeggini, Mark Daly, Bjørn Pasternak; **HUNT Research Centre:** Jørn Søberg Fenstad, Anne Jorunn Vikdal, Marit Næss; **HUNT Cloud:** Oddgeir L. Holmen, Sandor Zeestraten, Tom Erik Røberg; **Data applications and registry linkages:** Maiken E. Gabrielsen, Anne Heidi Skogholt; **Low-pass whole sequencing genome bioinformatics and statistical analysis:** He Zhang, Hyun Min Kang, Jin Chen; **Array genotyping:** Sten Even Erlandsen, Vidar Beisvåg; **GWAS bioinformatics, QC, imputation and statistical analysis:** Wei Zhou, Jonas Nielsen, Lars G. Fritsche, Hyun Min Kang, Oddgeir L. Holmen, Laurent Thomas and Ben M. Brumpton; **CNV calling:** Ellen Schmidt, Ryan Mills; **Statistical methods development for analyzing HUNT data:** Wei Zhou, Shawn Lee, Hyun Min Kang; **Communications management:** Bethany Klunder.

## Author Contributions

K.H., C.J.W and B.M.B wrote the first draft of the manuscript. W.Z., J.N., L.G.F., H.M.K., O.L.H., L.T and B.M.B contributed to analysis. K.H, C.JW., G.R.A., O.L.H and B.M.B contributed to funding acquisition. K.H., C.J.W., O.L.H., M.B., G.R.A, B.O.Å and B.M.B contributed to project administration. L.T., E.C., S.G., B.W. and B.M.B. contributed to visualization. All authors reviewed and edited the paper.

