## Supplementary Information for "The HUNT Study: a population-based cohort for genetic research"

### Supplementary Note

*Genotyping Array Design*

We aimed to identify as many high-quality genetic variants among HUNT participants as possible. Towards this aim, we developed a list of custom content for inclusion on one of four Illumina Human Core Exome arrays (HumanCoreExome12 v1.0, HumanCoreExome12 v1.1, UM HUNT Biobank v1.0 and UM HUNT Biobank v2.0) to directly genotype (i) 16,116 missense and loss-of-function variants as well as 1,072 lipid-associated variants identified from low-pass sequencing, (ii) 149 variants observed in Norwegian clinics for familial hypercholesterolemia, (iii) 5,324 Neanderthal variants, and (iv) 32,868 not-previously-observed variants predicted to introduce a premature stop codon in any of 56 genes in which protein-altering variants are deemed clinically actionable by The American College of Medical Genetics and Genomics (ACMG56)[1] **(Supplementary Table 4)**. Additionally, for the genotyping of HUNT4, we included variants for traits of interest including psoriasis, depression, alcohol use disorder, breast cancer, liver function, and bone mineral density; variants in the GWAS catalog; and loss-of-function variants available in TOPMed but poorly imputable in HUNT samples.

*Genotyping Procedures*

Protocols were carefully planned to mitigate any possible batch effects from the genotyping process. Sample assignments to plates and plate positions were randomized and sample sets that needed to be grouped together (e.g., based on robot requirements for liquid volume handling or a requirement for re-precipitation of DNA, etc.) were randomized within each subgroup. Within each plate, genetically determined sex was evaluated against expected sex to identify any plate orientation issues. To enable this during genotype calling, new HUNT-specific cluster files were developed for the genotyping arrays using GenomeStudio, which had to be specific to each array version. Following genotype calling, allele frequencies were examined between array versions and any variants that demonstrated significant association with batch or array versions were excluded. Limited manual validation of GenomeStudio calls (a few thousand variants) were performed. Quality control was performed based upon the approach developed by the Johns Hopkins Center for Inherited Disease Research (CIDR) and that of Guo et al[2].

In total, DNA from 90,582 were genotyped using one of four different Illumina HumanCoreExome arrays (HumanCoreExome12 v1.0, HumanCoreExome12 v1.1, UM HUNT Biobank v1.0 and UM HUNT Biobank v2.0). After a first round of automatic clustering in GenomeStudio (including samples with call rate > 95%), samples that failed to reach a 99% call rate, had contamination > 2.5% as estimated with BAF, large chromosomal copy number variants, lower call rate of a technical duplicate pair and twins, gonosomal constellations other than XX and XY, or whose inferred sex contradicted the reported gender, were excluded. Samples that passed quality control were analysed. Genomic position, strand orientation and the reference allele of genotyped variants were determined by aligning their probe sequences against the human genome (Genome Reference Consortium Human genome build 37 and revised Cambridge Reference Sequence of the human mitochondrial DNA; ) using BLAT[4]. Variants were excluded if (1) their probe sequences could not be perfectly mapped to the reference genome, cluster separation was < 0.3, Gentrain score was < 0.15, showed deviations from Hardy Weinberg equilibrium in unrelated samples of European ancestry with p-value < 0.0001), their call rate was < 99%, or another assay with higher call rate genotyped the same variant. Ancestry of all samples was inferred by projecting all genotyped samples into the space of the principal components of the Human Genome Diversity Project (HGDP) reference panel (938 unrelated individuals; downloaded from http://csg.sph.umich.edu/chaolong/LASER/)[5, 6]. For genotyping batches from HUNT2 and HUNT3, PLINK v1.90[7] was used and recent European ancestry was defined as samples that fell into an ellipsoid spanning exclusively European populations of the HGDP panel. For genotyping from HUNT4, we predicted ancestry using an online singular value decomposition and shrinkage adjustment algorithm (FRAPOSA) with the same reference panel[8]. The different arrays were harmonized by reducing to a set of overlapping variants and excluding variants that showed frequency differences > 15% between data sets, or that were monomorphic in one and had MAF > 1% in another data set. The resulting genotype data were phased using Eagle2 v2.3[9].http://genome.ucsc.edu) using BLAT[4]. Variants were excluded if (1) their probe sequences could not be perfectly mapped to the reference genome, cluster separation was < 0.3, Gentrain score was < 0.15, showed deviations from Hardy Weinberg equilibrium in unrelated samples of European ancestry with p-value < 0.0001), their call rate was < 99%, or another assay with higher call rate genotyped the same variant. Ancestry of all samples was inferred by projecting all genotyped samples into the space of the principal components of the Human Genome Diversity Project (HGDP) reference panel (938 unrelated individuals; downloaded from http://csg.sph.umich.edu/chaolong/LASER/)[5, 6]. For genotyping batches from HUNT2 and HUNT3, PLINK v1.90[7] was used and recent European ancestry was defined as samples that fell into an ellipsoid spanning exclusively European populations of the HGDP panel. For genotyping from HUNT4, we predicted ancestry using an online singular value decomposition and shrinkage adjustment algorithm (FRAPOSA) with the same reference panel[8]. The different arrays were harmonized by reducing to a set of overlapping variants and excluding variants that showed frequency differences > 15% between data sets, or that were monomorphic in one and had MAF > 1% in another data set. The resulting genotype data were phased using Eagle2 v2.3[9]. After quality control, 88,615 samples remained for further analysis.

*Imputation*

The imputation described here is limited to the 69,716 samples of recent European ancestry from HUNT2-3, as the work on HUNT4 is ongoing. Samples were imputed using Minimac3 (v2.0.1, http://genome.sph.umich.edu/wiki/Minimac3)[10] with default settings (2.5 Mb reference-based chunking with 500kb windows) and the HUNT-WGS customized Haplotype Reference consortium release 1.1 (HRC v1.1) for autosomal variants and HRC v1.1 for chromosome X variants[11]. The HUNT-WGS customized reference panel represented the merged panel of two reciprocally imputed reference panels: (1) 2,201 low- coverage (5x) whole-genome sequenced samples from the HUNT study (HUNT-WGS) and (2) HRC v1.1 with 1,023 overlapping HUNT WGS samples removed before merging. Since only 1,200 HUNT samples were sequenced at the time the HRC was established, we instead merged all HUNT-WGS samples (including indels) with the non-HUNT HRC samples to create a combined HRC and HUNT-WGAS imputation reference panel. Additionally, we recently performed imputation including samples of non-European ancestry (n=70,517) using the 60,039 TOPMed reference genomes using Minimac4 (v1.0).

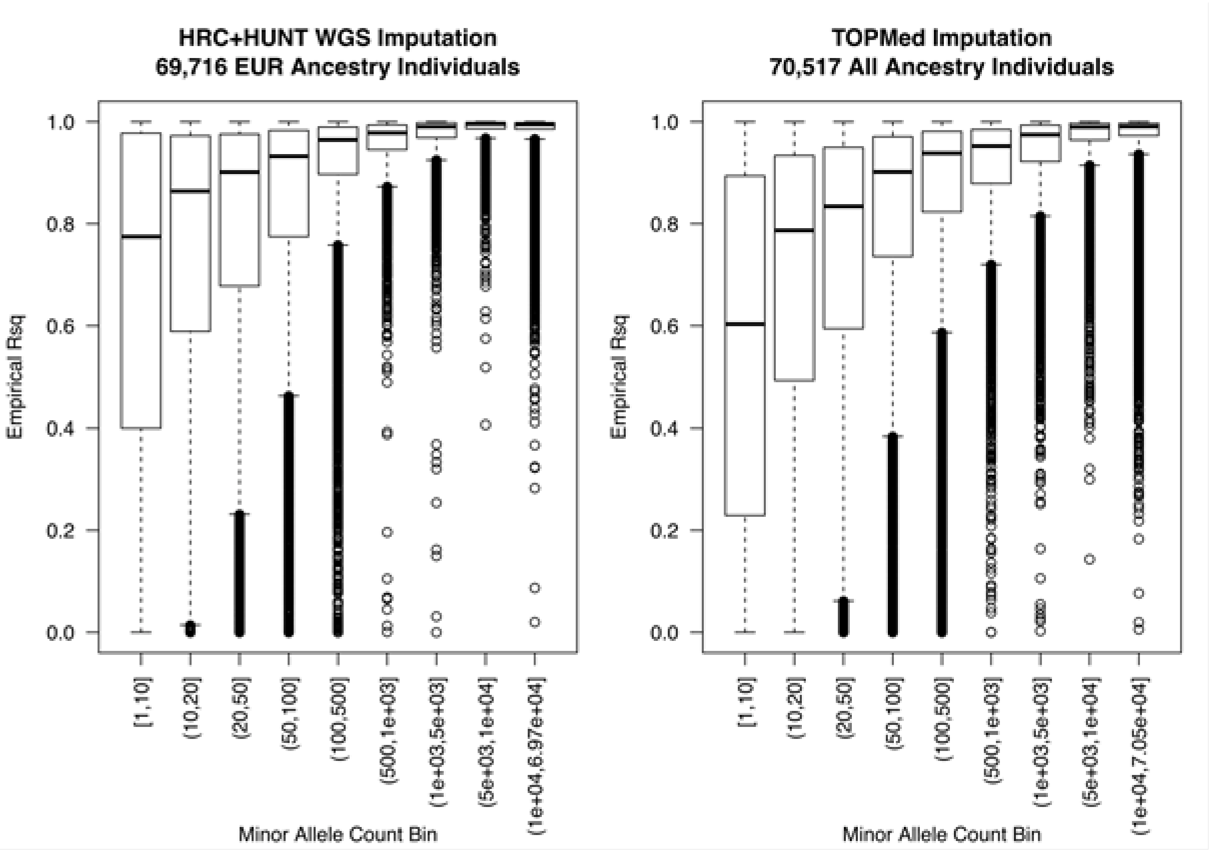


**Supplementary Figure 1: Imputation quality from HRC+HUNT WGS and TOPMed imputation panels in HUNT2-3**

HRC: Haplotype Reference Consortium, HUNT: Trøndelag Health Study, TOPMed: Trans-Omics for Precision Medicine, WGS: Whole genome sequencing

Reference panel: 1000 Genomes, HGDP SNPs of 2492 unrelated individuals

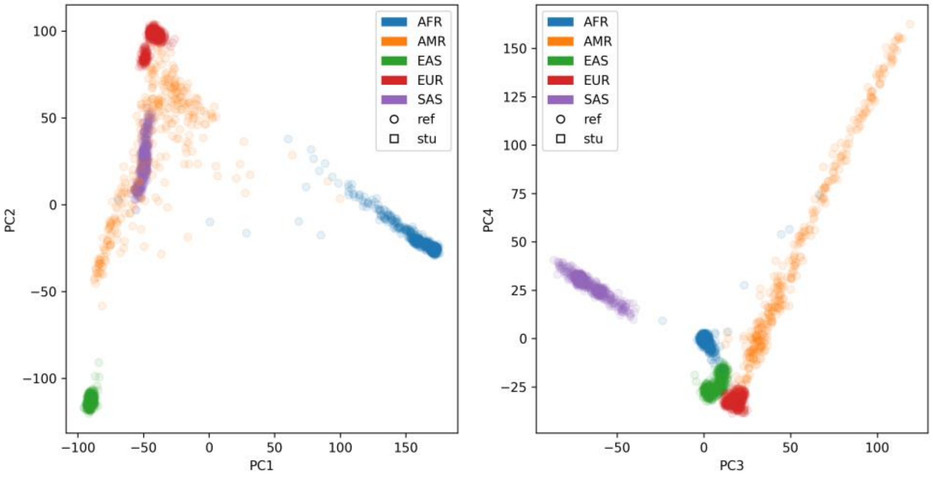


Study: HUNT2-4 samples

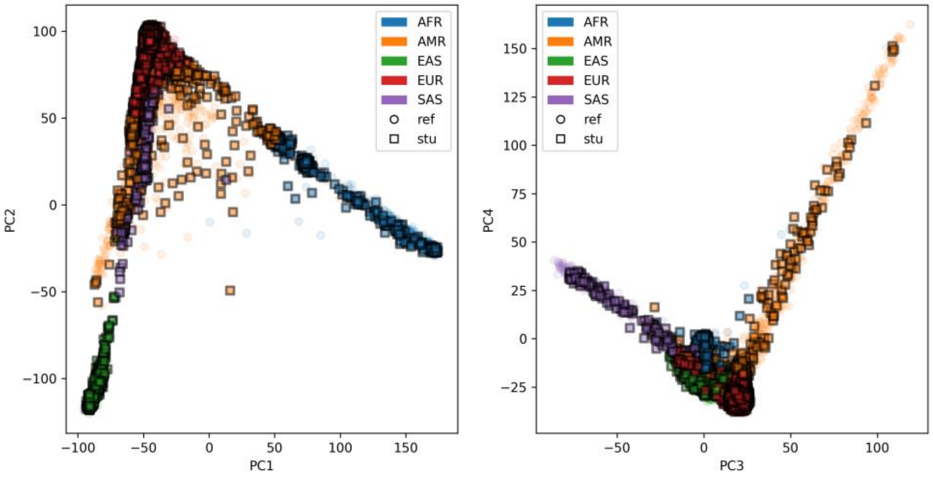


Study: HUNT2-4 samples of European ancestry

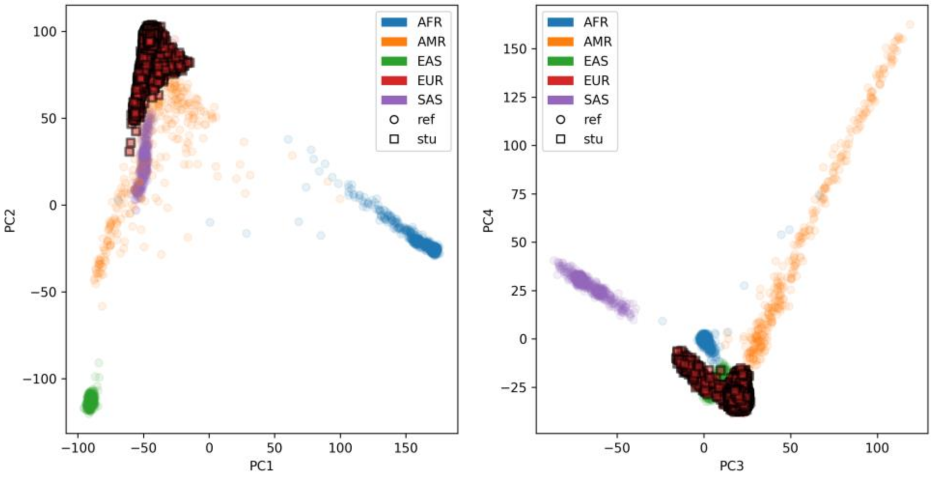

**Supplementary Figure 2: Plot of the first 4 principal components of ancestry projected with FRAPOSA onto the 1000 Genomes Project for the genotyped HUNT2-4 samples (N=88 615). 1,571 (<2%) samples with non-European ancestries were excluded from further genetic studies.**

HGDP: Human Genome Diversity Project, HUNT: Trøndelag Health Study, SNP: Single-nucleotide polymorphism.

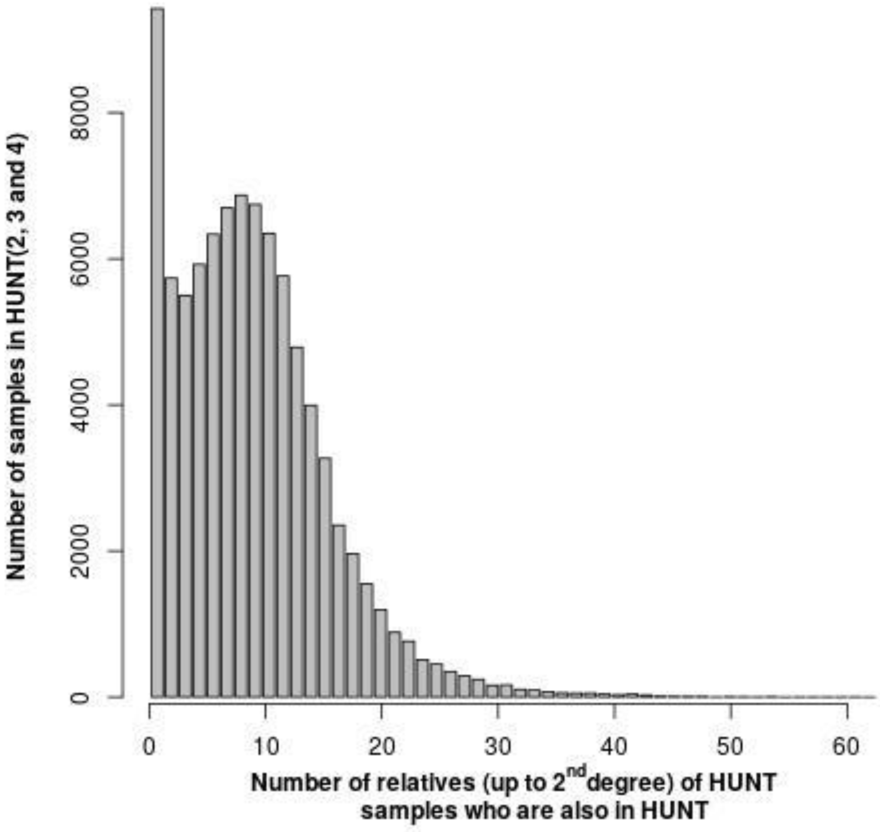


**Supplementary Figure 3: Histogram of numbers of up to 2nd degree relatives of HUNT2-4 samples who are also in HUNT (N=88 615).**

HUNT: Trøndelag Health Study

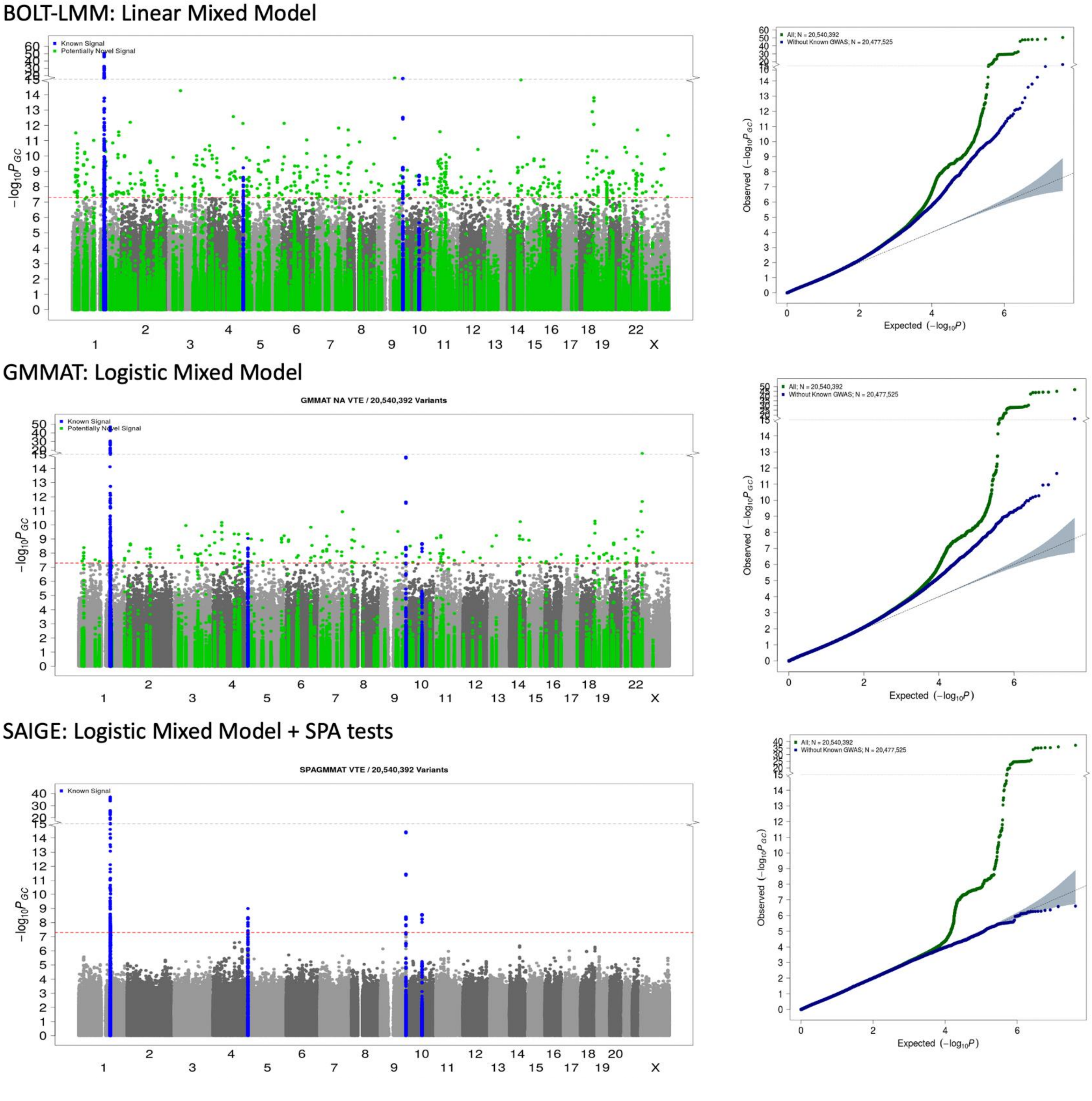


**Supplementary Figure 4: Analysis of venous thromboembolism (2 325 cases, 65 294 controls, case:control=0.036) in HUNT2-3 samples using BOLT-LMM, GMMAT and SAIGE (to account for relatedness and control for unbalanced case-control imbalance) of binary phenotypes.**

HUNT: Trøndelag Health Study

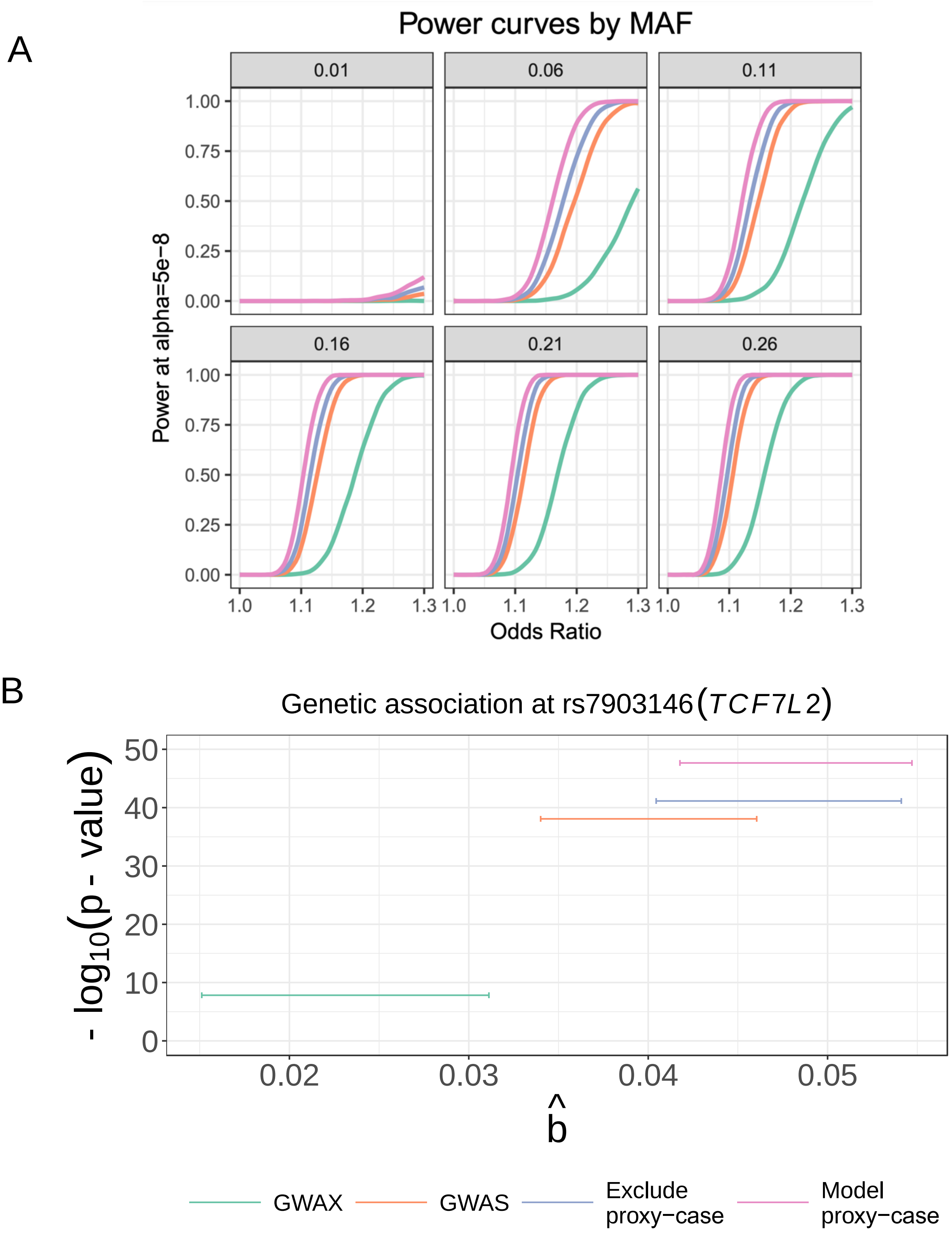


**Supplementary Figure 5: Power and empirical results for GWAS by proxy.**

**Panel A: Simulated power curves for proxy-case models.** Given a total biobank size of 100,000 for a disease prevalence of 10% and heritability of disease liability of 10% a biobank could have cases (n=10,000), proxy-cases (n=16,814) and controls (n=73,186). By performing linear regression with various methods of including proxy-cases, we can estimate the power at genome wide significance (p-value < 5e-8) across minor allele frequencies (0.01 to 0.26) and odds ratios (1 to 1.3). GWAX uses only proxy-cases as cases[12], GWAS is standard cases versus controls without identifying proxy-cases within the controls, excluding proxy-cases removes these for cleaner controls, and modelling proxy-cases uses coefficient of kinship for controls (F=0), proxy-cases (F=0.5), and cases (F=1) respectively. **Panel B: Association of the genetic variant rs7903146 with type 2 diabetes in HUNT across proxy-case models.** Empirical effect size and p-value for association at known type 2 diabetes variant rs7903146 in TCF7L2 in HUNT across methods of modelling proxy-cases. Linear mixed model as implemented in BOLT-LMM was used for 5.382 type 2 diabetes cases, 4,747 proxy-cases, and 20,284 controls. Proxy-cases were identified using self-reported first-degree family history of diabetes from HUNT questionnaires.

GWAS: Genome-wide association study, GWAX: Genome-wide association study by proxy, HUNT: Trøndelag Health Study, MAF: Minor allele frequency.

**Supplementary Table 1: Summary of the variants in the HUNT whole-genome sequencing reference panel containing 2,201 individuals with average sequencing depth 5x.**

| Variant Type | Total number of variants | Mean number of variants per individual (SD) | Mean number of unique variants per individual (SD) | % in 1000 Genomes | Number of novel variants* |
|---|---|---|---|---|---|
| **Splice** | 1,265 | 71.5(4.6) | 0.2(0.47) | 36.6 | 355 |
| **Nonsense** | 2,432 | 71.5(6) | 0.43(0.74) | 36.6 | 585 |
| **Missense** | 113,576 | 9,480(113) | 13.8(13.6) | 56.3 | 13,927 |
| **Synonymous** | 77,699 | 10,707(100) | 7.1(7.5) | 68.5 | 5,935 |
| **Noncoding** | 20,050,237 | 3,342,839(15,415) | 1,531(906) | 68.7 | 4,030,199 |
| **Total** | 20,245,209 | 3,363,168(15,522) | 1,552(919) | 68.6 | 4,051,001 |

*Novel: not reported in dbSNP 144[13], 1000 Genomes Phase 3[14], UK10K[15], ESP6500 (*W. NHLBI GO Exome Sequencing Project (ESP) Seattle*, 2013), or ExAC.r0.3[16]

ANNOVAR[17] was used for the annotation with the default setting. Splice variants are variants within 2-bp of a splicing junction.

HUNT: Trøndelag Health Study, SD: Standard deviation.

**Supplementary Table 2: Overview of mandatory national registries, other national and regional registries.**

| Register; Name in Norwegian (English name or description) | Type of register | Data owner; Name in Norwegian (English name or translation where an English name is not provided) | Data from or period of collection |
|---|---|---|---|
| Dødsårsaksregisteret (Cause of Death Registry) | National registry (mandatory) | Folkehelseinstituttet (Norwegian institute of public health) | 1951 |
| Kreftregisteret (Cancer Registry of Norway) | National registry (mandatory) | Oslo Universitetessykehus (Oslo University Hospital) | 1952 |
| Medisinsk fødselsregister (Medical Birth Registry of Norway) | National registry (mandatory) | Folkehelseinstituttet (Norwegian institute of public health) | 1967 |
| Meldingssytem for smittsomme sykdommer (MSIS) (Norwegian Surveillance System for Communicable Diseases) | National registry (mandatory) | Folkehelseinstituttet (Norwegian institute of public health) | 1977 |
| Nasjonalt vaksinasjonsregister (SYSVAK) (Norwegian Immunisation Registry) | National registry (mandatory) | Folkehelseinstituttet (Norwegian institute of public health) | 1976 |
| Forsvarets helseregister (Registry of the Norwegian Armed Forces Medical Services) | National registry (mandatory) | Forsvaret (Armed Forces Medical) | 2001 |
| Norsk pasientregister (Norwegian Patient Registry) | National registry (mandatory) | Helsedirektoratet (Norwegian Directorate of Health) | 1997 |
| Nasjonalt register over hjerte- og karlidelser (Norwegian Cardiovascular Disease Registry) | National registry (mandatory) | Folkehelseinstituttet (Norwegian institute of public health) | 2012 |
| System for bivirkningsrapportering (System for reporting of unwanted effects) | National registry (mandatory) | Legemiddelverket (Norwegian Medicines Agency) | 2008 |
| Kommunalt pasient- og brukerregister (Norwegian Registry of Primary Health Care) | National registry (mandatory) | Helsedirektoratet (Norwegian Directorate of Health) | 2017 |
| Helsearkivregisteret (Norwegian Medical Archives Registry) | National registry (mandatory) | Riksarkivaren (National Archives of Norway) | 2019 |
| Individbasert pleie- og omsorgsstatistikk (IPLOS) (Individual-based Statistics for Nursing and Care Services) | National registry (mandatory) | Helsedirektoratet (Norwegian Directorate of Health) | 2007-2017 |
| Norsk overvåkningssystem for antibbiotikabbruk og helsetjenesteassosierte infeksjoner (NOIS) (Norwegian Surveillance System for Antibiotic Use and Healthcare-Associated Infections) | National registry (mandatory) | Folkehelseinstituttet (Norwegian institute of public health) | 2005 |

| | | | |
|---|---|---|---|
| Norsk overvåkningssytem for antibiotikaresistens hos mikrober (NORM) (Norwegian Surveillance System for Antimicrobial Drug Resistance) | National registry (mandatory) | Folkehelseinstituttet (Norwegian institute of public health) | 1951 |
| Register over svangerskapsavbrudd (Abortregisteret) (Registry of Pregnancy Termination) | National registry (mandatory) | Folkehelseinstituttet (Norwegian institute of public health) | 1979 |
| Reseptbasert legemiddelregister (Norwegian Prescription Database) | National registry (mandatory) | Folkehelseinstituttet (Norwegian institute of public health) | 2004 |
| Resistensovervåkning av virus i Norge (RAVN) (Norwegian Surveillance System for Resistance Against Anti-virals) | National registry (mandatory) | Folkehelseinstituttet (Norwegian institute of public health) | 2013 |
| Norsk Hjerneslagregister (Norwegian Stroke Registry) | Medical quality registry | Folkehelseinstituttet (Norwegian institute of public health) | 2012 |
| Norsk diabetesregister for voksne (Norwegian Diabetes Register for Adults) | Medical quality registry | Helse Bergen (Bergen Hospital Trust) | 2006 |
| Norsk Kvalitets-og oppfølgingsregister for cereberal parese (NorCP) (Norwegian Quality and Follow-up Register for cereberal palsy) | Medical quality registry | Sykehuset i Vestfold (Vestfold Hospital Trust) | 1996 |
| Norsk Nyreregister (Norwegian Renal Registry) | Medical quality registry | Oslo Universitetessykehus (Oslo University Hospital) | 2016 |
| Nasjonalt medisinsk kvalitetregister for barne- og ungdomsdiabetes (Norwegian Childhood Diabetes Registry) | Medical quality registry | Oslo Universitetessykehus (Oslo University Hospital) | 1973 |
| Nasjonalt kvalitetsregister for prostatakreft (Norwegian Prostate Cancer Registry) | Medical quality registry | Kreftregisteret (Cancer registry of Norway) | 2004 |
| Norsk hjerteinfarktregister (Norwegian Myocardial Infarction Registry) | Medical quality registry | Folkehelseinstituttet (Norwegian institute of public health) | 2013 |
| Norsk hjertekirurgiregister (Norwegian Register for Cardiac Surgery) | Medical quality registry | Folkehelseinstituttet (Norwegian institute of public health) | 1994 |
| Norsk hjertesviktregister (Norwegian Heart Failure Registry) | Medical quality registry | Folkehelseinstituttet (Norwegian institute of public health) | 2012 |
| Norsk register for invasiv kardiologi (NORIC) (Norwegian Registry for Invasive Cardiology) | Medical quality registry | Folkehelseinstituttet (Norwegian institute of public health) | 2012 |
| Norsk hjertestansregister (Norwegian Cardiac Arrest Registry) | Medical quality registry | Folkehelseinstituttet (Norwegian institute of public health) | 2013 |
| Nasjonalt register for ablasjonsbehandling og elektrofysiologi i Norge (AblaNor) (Norwegian Ablation and Electrophysiology Registry) | Medical quality registry | Helse Bergen (Bergen Hospital Trust) | 2019 |
| Nasjonalt kvalitetsregister for tykk- og endetarmskreft (Norwegian Colorectal Cancer Registry) | Medical quality registry | Kreftregisteret (Cancer registry of Norway) | 1993 |

| | | | |
|---|---|---|---|
| Nasjonalt kvalitetsregister for brystkreft (Norwegian Breast Cancer Registry) | Medical quality registry | Kreftregisteret (Cancer registry of Norway) | 2009 |
| Nasjonalt kvalitetsregister for melanom (Norwegian Melanoma Registry) | Medical quality registry | Kreftregisteret (Cancer registry of Norway) | 2008 |
| Nasjonalt kvalitetsregister for gynekologisk kreft (Norwegian Gynecological Cancer Registry) | Medical quality registry | Kreftregisteret (Cancer registry of Norway) | 2012 |
| Nasjonalt kvalitetsregister for lungekreft (Norwegian Lung Cancer Registry) | Medical quality registry | Kreftregisteret (Cancer registry of Norway) | 2013 |
| Nasjonalt kvalitetsregister for lymfoide maligniteter (Norwegian Registry of Lymphoid Malignancies) | Medical quality registry | Kreftregisteret (Cancer registry of Norway) | 2013 |
| Nasjonalt register for langtids mekanisk ventilasjon (Norwegian Register for Long-Term Mechanical Ventilation) | Medical quality registry | Helse Bergen (Bergen Hospital Trust) | 2012 |
| Norsk MS-register og biobank (Norwegian MS-registry and Biobank) | Medical quality registry | Helse Bergen (Bergen Hospital Trust) | 2001 |
| Norsk register for arvelige og medfødte nevromuskulære sykdommer (Norwegian Registry for Hereditary and Congenital Neuromuscular Disorders) | Medical quality registry | Universitetssykehuset i Nord-Norge (University Hospital of North Norway HF) | 2008 |
| Norsk register for personer som utredes for kognitive symptomer i spesialisthelsetjenesten - NorKog (Norwegian Registry of Persons Assessed for Cognitive Symptoms in Specialist Health Care Services (NorCog)) | Medical quality registry | Oslo Universitetessykehus (Oslo University Hospital) | 2008 |
| Nasjonalt register for leddproteser (Norwegian Arthroplasty Register) | Medical quality registry | Helse Bergen (Bergen Hospital Trust) | 1987 |
| Nasjonalt hoftebruddregister (Norwegian Hip Fracture Register) | Medical quality registry | Helse Bergen (Bergen Hospital Trust) | 2005 |
| Nasjonalt korsbåndregister (Norwegian Knee Ligament Registry (NKLR)) | Medical quality registry | Helse Bergen (Bergen Hospital Trust) | 2004 |
| Nasjonalt kvalitetsregister for ryggkirurgi (NKR) (Norwegian Registry for Spine Surgery (NORspine)) | Medical quality registry | Universitetssykehuset i Nord-Norge (University Hospital of North Norway HF) | 2006 |
| Norsk nakke- og ryggregister (Norwegian neck and back register) | Medical quality registry | Universitetssykehuset i Nord-Norge (University Hospital of North Norway HF) | 2015 |
| Norsk kvalitetsregister for artrittsykdommer (NorArtritt) (Norwegian Arthritis Registry (NorArthritis) | Medical quality registry | Helse Bergen (Bergen Hospital Trust) | 2014 |
| Nasjonalt barnehofteregister (Norwegian Paediatric Hip Register) | Medical quality registry | Helse Bergen (Bergen Hospital Trust) | 2010 |
| Norsk register for analinkontinens (Norwegian Registry for Surgical Treatment of Anal Incontinence) | Medical quality registry | Universitetssykehuset i Nord-Norge (University Hospital of North Norway HF) | 2013 |

| | | | |
|---|---|---|---|
| Norsk ERCP-register (Gastronet) (Norwegian ERCP Registry) | Medical quality registry | Sykehuset Telemark (Telemark Hospital Trust) | 2003 |
| Norsk register for gastrokirurgi (NoRGast) (Norwegian Registry for Gastrointestinal Surgery (NoRGast)) | Medical quality registry | Universitetssykehuset i Nord-Norge (University Hospital of North Norway HF) | 2014 |
| Norsk gynekologisk endoskopiregister (Norwegian Gynecological Endoscopy Registry (NGER)) | Medical quality registry | Sykehuset i Vestfold (Vestfold Hospital Trust) | 2013 |
| Norsk kvinnelig inkontinensregister (NKIR) (Norwegian Female Incontinence Registry) | Medical quality registry | Oslo Universitetessykehus (Oslo University Hospital) | 1998 |
| Norsk intensiv- og pandemiregister (NIPaR) (Norwegian Intensive Care and Pandemic Registry (NIPaR)) | Medical quality registry | Helse Bergen (Bergen Hospital Trust) | 1998 |
| Norsk nyfødtmedisinsk kvalitetregister (Norwegian Neonatal Network database) | Medical quality registry | Folkehelseinstituttet (Norwegian institute of public health) | 2004 |
| Nasjonalt traumeregister (Norwegian Trauma Registry) | Medical quality registry | Oslo Universitetessykehus (Oslo University Hospital) | 2015 |
| Norsk ryggmargsskaderegister (NorSCIR) (Norwegian Registry for Spinal Cord Injury) | Medical quality registry | St. Olavs hospital (St. Olav's University Hospital) | 2011 |
| Nasjonalt register for organspesifikke autoimmune sykdommer (ROAS) (Norwegian Registry for Organ-Specific Autoimmune Diseases) | Medical quality registry | Helse Bergen (Bergen Hospital Trust) | 2006 |
| Nordisk register for hidradenitis suppurativa (HISREG) (Scandinavian Registry for Hidradenitis Suppurativa) | Medical quality registry | Universitetssykehuset i Nord-Norge (University Hospital of North Norway HF) | 2012 |
| Norsk kvalitetsregister for behandling av spiseforstyrrelser (NorSpis) (Norwegian Quality Registry for Eating Disorders ) | Medical quality registry | Nordlandsykehuset (Nordland Hospital Trust) | 2017 |
| Nasjonalt Kvalitetsregister for smertebehandling (SmerteReg) (Norwegian Acute Pain Service (APS) Registry) | Medical quality registry | Helse Bergen (Bergen Hospital Trust) | 2014 |
| Norsk kvalitetsregister for leppe-kjeve-ganespalte (LKG-registeret) (Norwegian Registry of Cleft Lip and Palate) | Medical quality registry | Helse Bergen (Bergen Hospital Trust) | 2011 |
| Norsk porfyriregister (Norwegian Porphyria Registry) | Medical quality registry | Helse Bergen (Bergen Hospital Trust) | 2002 |
| Norsk kvalitetsregister for fedmekirurgi (SOReg-N) (Scandinavian Obesity Surgery Registry Norway) | Medical quality registry | Helse Bergen (Bergen Hospital Trust) | 2015 |
| Norsk karkirurgisk register (NORKAR) (Norwegian Vascular Surgery Registry) | Medical quality registry | Folkehelseinstituttet (Norwegian institute of public health) | 2015 |
| Nasjonalt kvalitetsregister for barnekreft (Norwegian Childhood Cancer Registry) | Medical quality registry | Kreftregisteret (Cancer registry of Norway) | 1985 |

| | | | |
|---|---|---|---|
| Norsk Parkinsonregister og biobank (Norwegian Parkinsons Registry and Biobank) | Medical quality registry | Stavanger universitetssjukehus | 2016 |
| Norsk vaskulittregister & biobank (NorVas) (Norwegian Vasculitis Registry and Biobank) | Medical quality registry | Universitetssykehuset i Nord-Norge (University Hospital of North Norway HF) | 2014 |
| Norsk Tonsilleregister (Norwegian Tonsil Surgery Register) | Medical quality registry | St. Olavs hospital (St. Olav's University Hospital) | 2017 |
| Nasjonalt kvalitetsregister for behandling av skadelig bruk eller avhengighet av rusmidler (KVARUS) (Norwegian Quality Register for Substance Abuse Treatment) | Medical quality registry | Stavanger universitetssjukehus (Stavanger University Hospital) | 2018 |
| Kommunehelsa Statistikkbank (Public health statistics on municipality level) | Statistics | Folkehelseinstituttet (Norwegian institute of public health) | 2012* |
| Norgeshelsa Statistikkbank (Public health statistics on county/national level) | Statistics | Folkehelseinstituttet (Norwegian institute of public health) | 2000* |
| A-ordningen (a coordinated service used by employers to report information about income and employees to NAV, Statistics Norway and the Norwegian Tax Administration) | Administrative | Statistisk sentralbyrå (Statistics Norway) (SSB) | 2015 |
| Arbeidsmarked (Labour market and earnings) | Administrative | Statistisk sentralbyrå (Statistics Norway) (SSB) | 1983-2014 |
| Barnevern (Child welfare) | Administrative | Statistisk sentralbyrå (Statistics Norway) (SSB) | 1993 |
| Befolkning (Population) | Administrative | Statistisk sentralbyrå (Statistics Norway) (SSB) | 1964 |
| Boforhold (Housing Conditions) | Administrative | Statistisk sentralbyrå (Statistics Norway) (SSB) | 2015 |
| Fastlegeregisteret (Norwegian GP Registry) | Administrative | Helsedirektoratet (Norwegian Directorate of Health) | 2001 |
| Folke- og boligtellinger (Population and Housing Census) | Administrative | Statistisk sentralbyrå (Statistics Norway) (SSB) | 1960 |
| Inntekt og forbruk (Income and consumption) | Administrative | Statistisk sentralbyrå (Statistics Norway) (SSB) | 1993 |
| Introduksjonsordningen (Introduction programme for immigrants) | Administrative | Statistisk sentralbyrå (Statistics Norway) (SSB) | 2005 |
| Kontantstøtte (cash-for-care benefits for parents with child(ren) between the ages 1 and 2 years who does not attend kindergarten full time) | Administrative | Statistisk sentralbyrå (Statistics Norway) (SSB) | 1999 |
| Kontroll og utbetaling av helserefusjoner (KUHR) (Norwegian Control and Payment of Health Reimbursements Database) | Administrative | Helsedirektoratet (Norwegian Directorate of Health) | 2006 |

| | | | |
|---|---|---|---|
| Lønn (Salary) | Administrative | Statistisk sentralbyrå (Statistics Norway) (SSB) | 1997 |
| Trygd (Benefits) | Administrative | Statistisk sentralbyrå (Statistics Norway) (SSB) | 1992 |
| Utdanning (Education) | Administrative | Statistisk sentralbyrå (Statistics Norway) (SSB) | 1970 |

*Started in 2012, but data is derived from different sources and therefore dates may vary.

**Supplementary Table 3. Relationship inference of genotyped samples from HUNT2-4 (N=88 721)*.**

| **Inference** | **Monozygotic twins** | **Parent-offspring** | **Full-siblings** | **2nd degree** |
|---|---|---|---|---|
| **Number of pairs** | 106 | 68124 | 44777 | 192318 |

*To provide a full overview of the relationships in HUNT we included 106 pairs of twins in this analysis. The twin with the highest genotyping call rate is kept for other analyses.

HUNT: Trøndelag Health Study

**Supplemental Table 4: Selection of variants for inclusion in custom content for the HUNT-Michigan Illumina Infinium Human CoreExome arrays.**

| **Selection criterion for custom content on the HUNT human core exome v1.0** | **Variants that passed DesignScore > 0.5 AND designed into array as custom content** | **Polymorphic variants in ~60,000 Norwegian individuals** |
|---|---|---|
| Identified from HUNT sequencing* AND missense and minor allele count = 1 AND observed in 12,000 ESP samples (https://genome.sph.umich.edu/wiki/Exome_Chip_Design) | 5427 | 4843 |
| Identified from HUNT sequencing* AND missense and minor allele count >= 2 | 9853 | 8941 |
| Identified from HUNT sequencing* AND LoF and minor allele count >= 1 | 836 | 731 |
| Identified from HUNT sequencing* AND p < 1x10-4 for lipids or MI | 1072 | 1006 |
| NHGRI GWAS catalog 5/21/2014 AND $p < 5x10^{-8}$ | 1139 | 1056 |
| GIANT associated variants NOT in NHGRI GWAS catalog | 246 | 184 |
| 96 candidate genes including ACMG56 – any codon that would theoretically exist to produce a LoF | 32,868* | 2548 ** |
| Norwegian LDLR mutations from FH clinic | 149 | 34 |
| Total custom variants | 51590 | 19343 |
| **Additional selection criterion for custom content on the HUNT human core exome v2.0** | **Variants that passed DesignScore > 0.5 AND designed into array as custom content** | **Polymorphic variants in 18537 Norwegian individuals** |
| LoF variant poorly imputed from TOPMed | 449 | 350 |
| Associated with bone mineral density | 3 | 3 |
| Associated with depression and/or alcohol use disorder | 20 | 20 |

| | | |
|---|---|---|
| Associated with liver function | 7 | 6 |
| Associated with psoriasis | 10 | 8 |
| Breast cancer risk variant | 204 | 123 |
| GWAS catalog variant | 1692 | 760 |
| Subtotal additional custom variants | 2385 | 1270 |
| **Total custom variants** | 53975 | 20613 |

* 32,868 putative LoF variants were assayed with only one bead type. For 21,640 out of 32,868 putative LoF variants, the alternate allele that would cause a premature stop codon cannot be distinguished from a missense or synonymous change.

ESP: Exome sequencing project, FH: Familial hypercholesterolemia, GWAS: Genome-wide association study, HUNT: Trøndelag Health Study, LDLR: Low density lipoprotein receptor, LoF: Loss-of-function, MI: Myocardial infarction, NHGRI: National Human Genome Research Institute, TOPMed: Trans-Omics for Precision Medicine.
